# Parametric analysis of early data on COVID-19 expansion in selected European countries

**DOI:** 10.1101/2020.03.31.20049155

**Authors:** Martin Spousta

## Abstract

We analyze the early data on COVID-19 expansion in selected European countries using an analytical parametric model. A description of the time dependence of the disease expansion and a method to evaluate trends of the expansion are proposed. Several features are observed in the data, namely a high predictability of the expansion of disease in Italy and a convergence of the “pushback” parameter towards a limiting value in all the countries where restrictive measures have been adopted. Basic predictions for the evolution of the disease expansion are made for selected countries with a stable evolution in the parametric space of the model. The findings presented here should contribute to the understanding of the behavior of the disease expansion and the role of the restrictive measures on the evolution of the expansion.

## 1. Introduction

The outbreak of new coronavirus SARS-CoV-2 causing severe respiratory tract infection in humans, known as COVID-19, is a global health concern. Restrictive measures have been adopted in many countries in order to mitigate the impact of the spread of the disease on public health system [1]. In this paper we propose a straightforward analytic description of the time dependence of the disease expansion under the restrictive measures and a method allowing to identify trends in the expansion and make predictions. The analytic parametric modeling can be used as an alternative to complex models for the disease expansion such as those reviewed e.g. in Ref. [2].

In the first section of this paper, we introduce the model, analyze the data which are available as of 31^*st*^ of March 2020 from selected European countries and discuss several features seen in the data. In the second section, we explain calculations for predictions and we make predictions for three countries which appear to be on a predictable trajectory in the parametric space of the model. Data used in this work are taken from [3], cross-checked using information system described in Ref. [4], and from [5].

## 2. Analytic parametric model of COVID-19 expansion

Number of newly infected people from one infected person has a power-law dependence on time. In general, the total number of infected people in time *t* can be fully characterized by a function

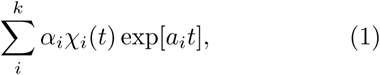

where *α*_*i*_ and *a*_*i*_ are unknown parameters. The sum goes trough individual centers of the disease expansion. These centers carry constant parameters *α*_*i*_ and *a*_*i*_ only over a limited time interval which is characterized by a step function *χ*_*i*_(*t*). The full description of the outbreak by a function (1) can be replaced by exponential dependence with time dependent parameter *a*(*t*), that is by the function

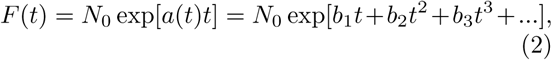

where the right-hand side of (2) represents a Taylor expansion of the time dependent parameter *a*. If one assumes that *a*(*t*) is constant or monotonically decreasing with time, one may assume that |*b*_*i*_| *>* |*b*_*i*+1_| and higher order coefficients of the Taylor expansion will contribute less to *F* (*t*). One may therefore start the description of the total number of infected people at a given time *t* with three approximations,

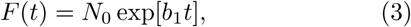

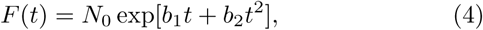

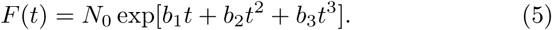

Approximation (3) is a pure exponential distribution which we shall observe if no human measures are taken to control the spread of the disease and no immunization of the population is assumed. Approximation (4) is modified exponential expansion where parameter *b*_2_, if negative, characterizes the “pushback” resulting from human measures to control the disease. Approximation (5) allows to control the validity of |*b*_*i*_| *>* |*b*_*i*+1_| assumption and a need for the higher order terms to model the spread of the disease.

Data can be fitted by (3), (4), and (5). The quality of the fits then decides which of the approximation serves as the best description of the data. When fitting the data over the full available time range from three representative European countries, namely Italy, France, and Czechia, we found that indeed |*b*_1_|≫| *b*_2_|≫| *b*_3_| and that approximation (5) has slightly worse *χ*^2^/NDOF than approximation (4) (typically by 10%). We may therefore conclude that approximation (5) and higher order terms are not needed for parameterizing the data.

To evaluate the evolution of the spread of the disease as a function of time, one can study the time dependence of parameters *b*_1_, *b*_2_, and *N*_0_. This can be done by fitting the data in time windows or by fitting the data between the time of the beginning and a given time *t*. The former method is more susceptible to fluctuations of the data which may occur within the time windows while the latter method may be biased by the change of the parameters in time. We tested both of these methods as well as various sizes of the time window with the conclusion that fitting from the beginning leads to less fluctuating values of parameters which exhibit the same trends and tend to converge to the same values as the values of parameters analyzed by fits in time windows. From now on, we therefore evaluate the evolution of parameters *b*_1_, *b*_2_, and *N*_0_ by fitting from the beginning up to a time *t* indicated on the *x*-axis of plots. The evolution of parameters *b*_1_, *b*_2_, and *N*_0_ is shown in upper left and lower panels of Figure 1. One can see that parameters are highly oscillating at the beginning but then for times greater than 20 days since the first registered cases, one can see a hint of a convergence. Parameter *b*_1_ first oscillates and then decreases and at later time it tends to converge for Italy and France towards a value in the interval of 0.3 *−* 0.35. Parameter *b*_2_ typically starts at zero which indicates the purely exponential spread of the disease at the beginning when no measures are taken. At later times, *b*_2_ tends to certain negative values. The study of correlations among parameters shows that the correlations are very high and the correlation coefficient may achieve values greater than 0.95 for certain times.

**Figure 1:**
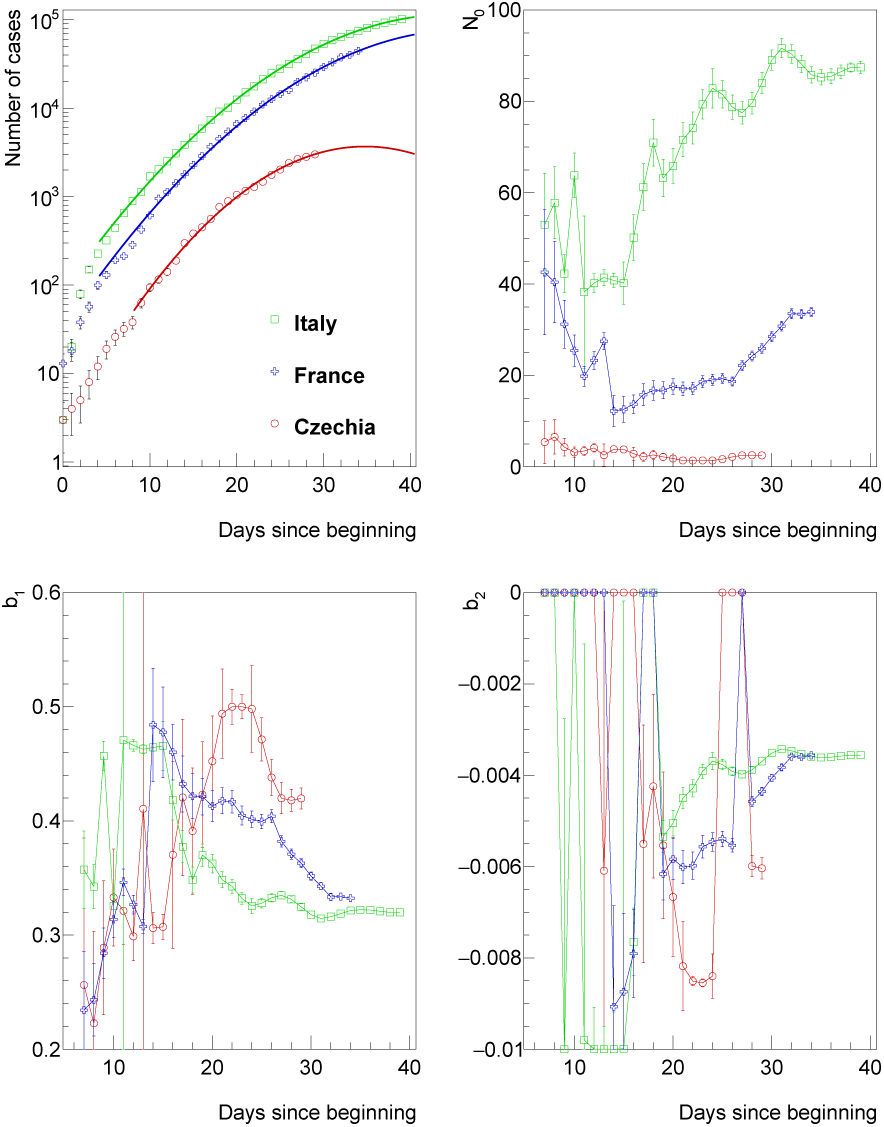
*Upper left:* Number of COVID-19 cases as a function of time since the first detection in a given country. The distribution is fitted by function defined in Equation (4). Fits ignore data from first five days and data with number of cases smaller than 100. Parameters of the fit: *N*_0_ *(upper right), b*_1_ *(lower left)*, and *b*_2_ *(lower right)* evaluated as a function of time since the first detection in a given country (i.e. fit stops at the time indicated on *x*-axis).

The parameter *b*_1_ can be interpreted as a parameter quantifying the uncontrolled spread of the disease. As such, it will be assumed that it is universal for all the European countries since they share approximately the same default density of interactions among inhabitants (cities of similar density, similar level of public transportation, similar default cultural behavior). Consequently, for subsequent studies, the parameter *b*_1_ was fixed to a value 0.32 representing a value preferred by a minimum *χ*^2^ condition for the fit in the country with the most stable evolution which is Italy.

The time evolution of *N*_0_ and *b*_2_ parameters in the case of fixed *b*_1_ is shown in Figure 2. One may identify several features seen from the behavior of parameters:

**Figure 2:**
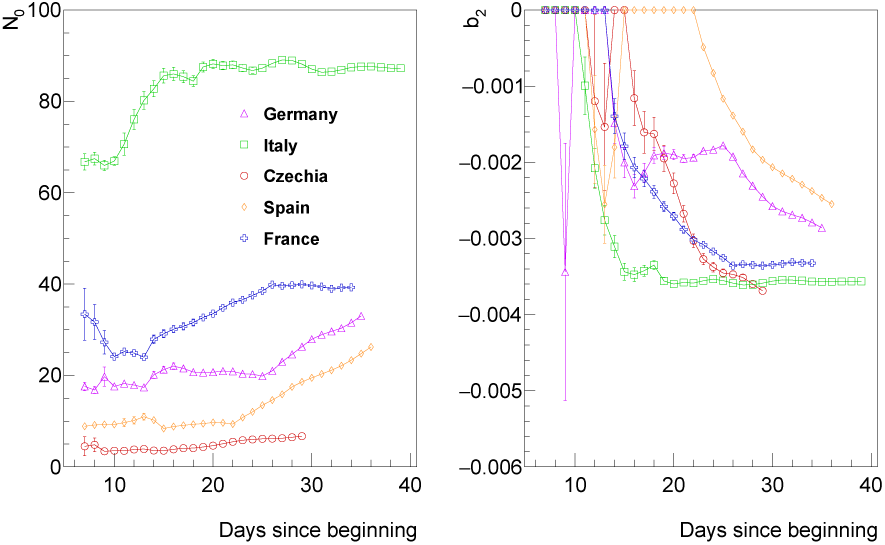
Parameters of the fit of the distribution of number of COVID-19 cases by function defined in Equation (4) with *b*_1_ parameter fixed. *N*_0_ *(left)* and *b*_2_ *(right)* evaluated as a function of time since the first detection in a given country (i.e. fit stops at the time indicated on *x*-axis).

1. Parameter *b*_2_ starts from zero (no restrictive measures), then decreases as expected for all the countries, and then it becomes constant for countries where restrictive measures are in place for a longer time (namely Italy and France).
2. All the countries seem to tend to the same value of the parameter *b*_2_.
3. Parameters *b*_2_ and *N*_0_ are consistent with a constant over the last *∼* 20 days in Italy. This means that the situation in Italy is highly predictable as further discussed in Section 3.
4. Parameter *N*_0_ is much higher for the case of Italy than for other countries.

The features 1*−* 3, if confirmed by other studies, open questions for a detailed epidemiological analysis: What is the reason for the existence of a limiting efficiency of applied measures which is indicated by the data? Is it e.g. due to the high-efficiency for the disease to spread inside closed communities? Or is it e.g. due to having a certain fraction of the society failing to follow correctly the measures? The analysis presented here obviously cannot answer these questions. At the same time, it allows us to assess the efficiency of applying the existing measures. For example, if we observe that the parameter *b*_2_ achieves a limiting value of Italy even in countries where more restrictive measures have been applied, such as mandatory usage of masks in Czechia, then this indicates that the use of these measures does not bring further reduction of the spread of the disease^1^.

Since *N*_0_ reflects the number of cases at the time zero, feature 4 implies that the infection was likely present in Italy even before the time zero. This may also explain the excessive values of parameter *b*_1_ observed for first *∼* 5 days.

We should note that when fixing *b*_1_, the values of parameters *N*_0_ and *b*_2_ still remain correlated. In some cases the correlation coefficient is greater than 0.9. This may limit the predictive power of the model and a straightforward interpretation of the parameter *N*_0_ as the number of cases at time zero. A way to reduce these correlations needs to be further studied. So far we have tested alternatives to (3)–(5), e.g.

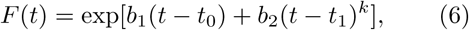

and

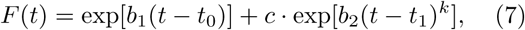

with *b*_*i*_, *t*_*i*_, and *c* being free parameters and *k* = 1, 2. None of these more ad-hoc functional forms provided a better description of the data than (4).

While *N*_0_ and *b*_2_ remain correlated, one can notice that the values of the *N*_0_ and *b*_2_ start to be constant in time for Italy and France even if the number of cases evolve with time. This builds a confidence in the predictive power of the model which is discussed in the next section.

## 3. Predicting further evolution

When being in the situation where all the parameters tend to constant values, one can predict the behavior of the spread of the disease using simple analytic formulae. An estimate of the time when the restrictive measures (*b*_2_) outperform the uncontrolled spread (*b*_1_), *t*_max_, that is the time when no new cases of the disease should be registered is given by a derivative of (4) with respect to time,

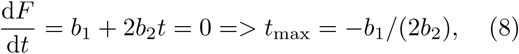

which also allows to calculate the total number of cases,

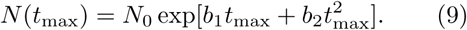

An estimate of the time when the number of new cases should stop to grow, *t*_Δ_, is an inflection point of *F* (*t*) and it is given by a second derivative of (4) with respect to time,

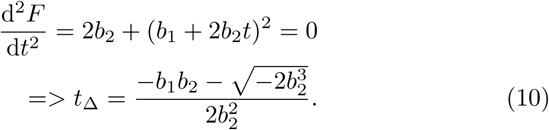

The results of the predictions using Equations (8)–(10) are shown in Figure 3 for countries with constant parameters of the fit which is Italy and France, and for Czechia which seems to be converging to a constant value of parameters as well. We do not attempt to introduce systematic uncertainties on the predictions at this point. Instead, we use evaluation of the distribution using two methods: using fits in time windows of seven days and the fits over the full distribution. As already mentioned before, the later method brings results less susceptible to fluctuations.

**Figure 3:**
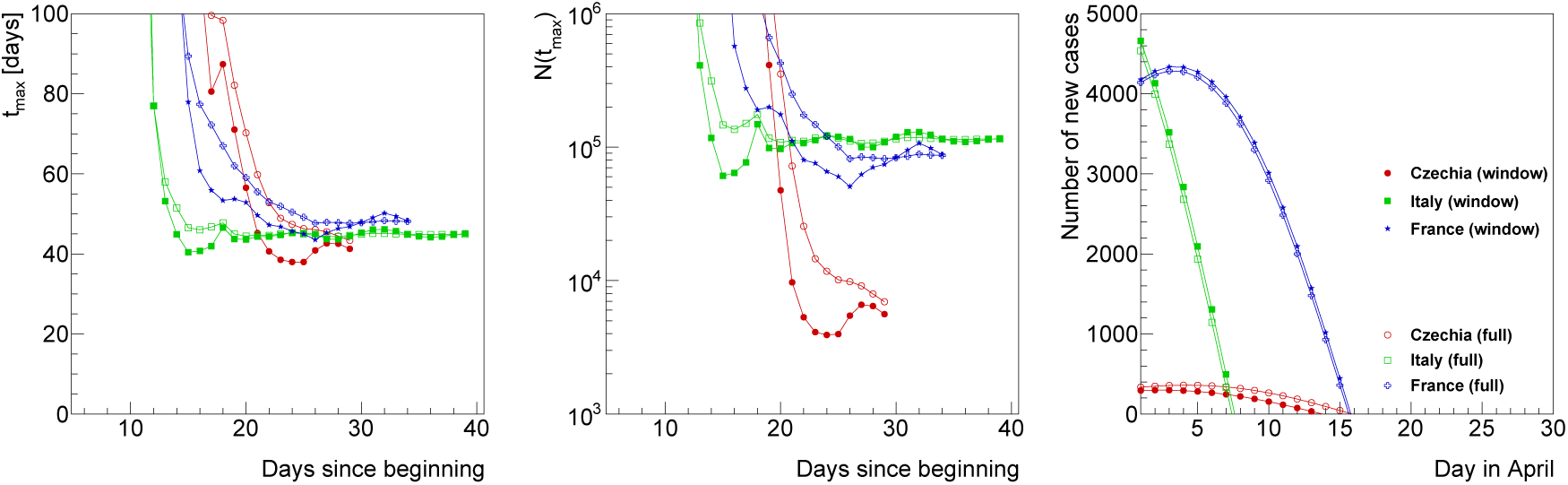
*Left:* Estimates of time when no new cases should occur, *t*_max_. *Middle:* total number of infected people at *t*_max_, *N* (*t*_max_). Both *t*_max_ and *N* (*t*_max_) are shown as a function of the last day since the beginning of the spread for which the data were included to the fit. *Right:* Prediction for the number of new cases as a function of day in April 2020.

Left and middle panel of Figure 3 show the prediction for the *t*_max_ values and *N* (*t*_max_) values, respectively, as a function of time since the first registered case. One can therefore see what prediction one would make in the past. The current prediction is represented by the last point of the distribution. One can again see that the prediction does not change over the last *∼*20 and *∼* 10 days in the case of Italy and France, respectively. As said before, these countries seem to be on a highly predictable trajectory which can be illustrated for Italy by evaluating the prediction for the total number of cases for today which one would make 20 days ago. The prediction would be 100970 cases while data for today (30th of March) show 101740 cases. For the case of Czechia, one can see an evolution towards more optimistic scenario as the time increases. At the same time, the prediction remains relatively stable over the last few days.

The prediction for the *t*_Δ_ is given in terms of the distribution of newly indicated cases as a function of time from today. One can see that the maximum of new cases should already been achieved in Italy, while in France and Czech Republic it is expected in about 4 and 1 *−* 4 days, respectively.

## 4. Summary and conclusions

We proposed a straightforward method for evaluating the temporal dependence of spread of COVID-19 disease. The analysis of the data indicate several features, namely the high predictability of the expansion of disease in Italy and a convergence of the “pushback” parameter towards a limiting value in all the countries where restrictive measures are applied. Predictions of the evolution of the spread of the disease are made for three countries that appear to be on a predictable trajectory in the parametric space of the model, namely for Italy, France, and Czechia.

The proposed model and analysis method represent an alternative to complex modeling which is simple to implement and independently verify, and which can be quickly extended towards more complete description of the situation in many different countries affected by the COVID-19 outbreak.

## Data Availability

This works uses external data available from links included in the work or below

https://onemocneni-aktualne.mzcr.cz/covid-19

https://www.worldometers.info/coronavirus/

## Acknowledgment

I’d like to thank Jiří Dolejší for useful discussions and suggestions and for careful reading the manuscript.

## Footnotes

1 1At the same time it would obviously not prove that this particular measure is not very efficient in the case when no other measures are taken. Additionally, if the *b*_2_ value for Czechia goes below the *b*_2_ value for Italy, it would indicate that more restrictive measures applied in Czechia may increase the efficiency in reducing the spread of the disease.

